# Association of Genetically Predicted Levels of Circulating Blood Lipids with Coronary Artery Disease Incidence

**DOI:** 10.1101/2024.04.23.24306257

**Authors:** Hasanga D. Manikpurage, Jasmin Ricard, Ursula Houessou, Jérôme Bourgault, Eloi Gagnon, Emilie Gobeil, Arnaud Girard, Zhonglin Li, Aida Eslami, Patrick Mathieu, Yohan Bossé, Benoit J. Arsenault, Sébastien Thériault

**Author notes:** <u>Address for correspondence</u> Sébastien Thériault, MD, MSc, FRCPC, Institut Universitaire de Cardiologie et de Pneumologie de Québec – Université Laval C-2113, 2725 chemin Ste-Foy, Québec (QC), Canada, G1V 4G5, T : - E.

## Abstract

**Background:** Estimating the genetic risk of coronary artery disease (CAD) is now possible by aggregating data from genome-wide association studies (GWAS) into polygenic risk scores (PRS). Combining multiple PRS for specific circulating blood lipids could improve risk prediction. Here, we sought to evaluate the performance of PRS derived from CAD and blood lipids GWAS to predict the incidence of CAD.

**Methods:** This study included individuals aged between 40 and 69 recruited in UK Biobank (UKB). We conducted GWAS for blood lipids measured by nuclear magnetic resonance in individuals without lipid-lowering treatments (n=73,915). Summary statistics were used to derive and calculate PRS in the remaining participants (n=318,051). A PRS_CAD_ was also derived using the CARDIoGRAMplusC4D GWAS. Hazard ratios (HR) for CAD (9,017 / 301,576; median follow-up time: 12.6 years) were calculated per standard deviation increase in each PRS. Discrimination capacity and goodness of fit of the models were evaluated.

**Results:** Out of 30 PRS, 28 were significantly associated with the incidence of CAD (*P*<0.05). The optimal combination of PRS included PRS for CAD, VLDL-C, total cholesterol and triglycerides. Discriminative capacities were significantly increased in the model including PRS_CAD_ and clinical risk factors (CRF) (C-statistic=0.778 [0.773-0.782]) compared to the model with CRF only (C-statistic=0.755 [0.751-0.760]). Although the C-statistic remained similar when independent lipids PRS were added to the model with PRS_CAD_ and CRF (C-statistic=0.778 [0.773-0.783]), the goodness of fit was significantly increased (chi-square test statistic=20.18, *P*=1.56e-04).

**Conclusions:** Although independently associated with CAD incidence, blood lipids PRS provide modest improvement in the predictive performance when added to PRS_CAD_.

**Highlights:**

- Genome-wide association studies were conducted on 29 selected lipid traits measured by nuclear magnetic resonance spectroscopy in 73,915 participants from UK Biobank who were not taking lipid-lowering treatment.
- Polygenic risk scores for 27 out of 29 of these traits were associated with the incidence of coronary artery disease (CAD) in 9,017 cases out of 301,576 individuals followed for a median of 12.6 years.
- When combined to a PRS for coronary artery disease, there was a significant but modest improvement in the discrimination capacity for incident CAD.
- PRS for certain lipid traits might help to stratify the risk of CAD.

**Graphical Abstract:** 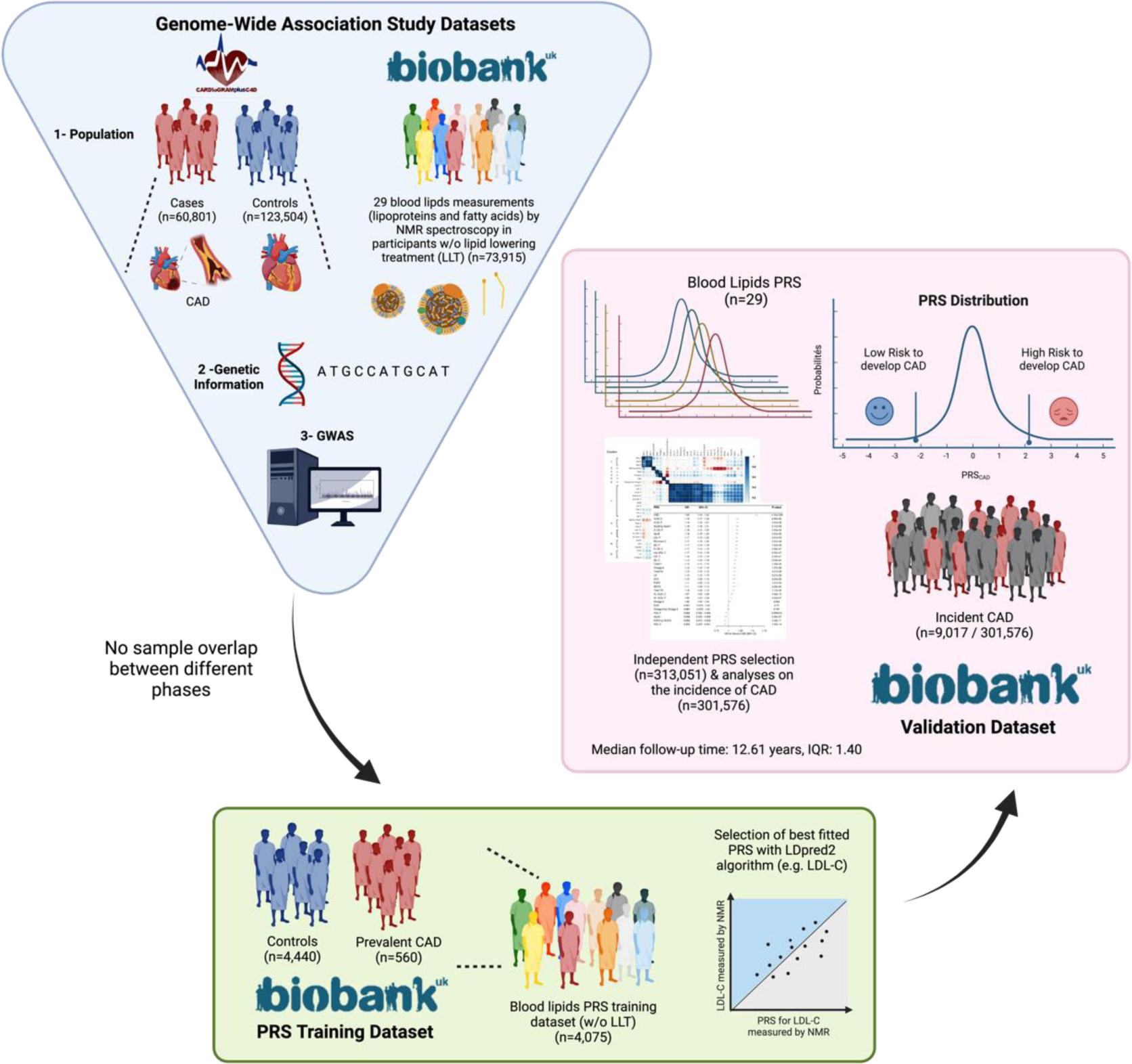

## Introduction

Circulating lipoproteins are recognized primordial risk factors for atherosclerotic cardiovascular diseases, such as coronary artery disease (CAD). The cholesterol content of low-density lipoproteins (LDL-C) is associated with atheroma development and progression and is routinely monitored for clinical management (1). Other lipoproteins have now been associated with CAD (2), including intermediate-density lipoproteins (IDL), very low-density lipoproteins (VLDL) and chylomicrons (also called extra-large very low-density lipoproteins, XL-VLDL) (3). Beyond cholesterol content, density, size, and number of particles of these lipoproteins have been identified as important determinants of atherosclerosis development and cardiovascular outcomes (2,4–6). The measurement of apolipoprotein B (ApoB) allows to estimate particle concentration of these atherogenic lipoproteins. Characteristics of these lipoproteins can be quantified using high-resolution methods such as nuclear magnetic resonance (NMR) spectroscopy (7). A recent study reported significant associations between such measurements and the incidence of major adverse cardiovascular events (8).

Circulating fatty acids (FA) have also been implicated in cardiovascular risk. Many are precursors of bioactive lipids involved in various biological mechanisms such as inflammatory processes or their resolution, immunomodulation, regulation of vascular tone, coagulation and cell migration (9). Recent results of the randomized clinical trial PREDIMED (*Prevención con Dieta Mediterránea*), in which adherence to a Mediterranean diet rich in omega-3 polyunsaturated fatty acids (PUFA) was compared to a low-fat diet, suggested a cardioprotective role of omega-3 PUFA by reducing the incidence of primary major cardiovascular events in high-risk individuals (10). Moreover, high doses of omega-3 PUFA-derived components led to a decrease in ischemic events in other randomized clinical trials, mainly in patients with hypertriglyceridemia (11,12).

Many factors influence blood lipid levels. Their intestinal absorption and downstream catabolism involve various enzymatic reactions and molecular processes. Each biological step is influenced by genetic variations, translating, for instance, in variations of enzyme or protein expression, structure or activity. This is supported by a recent meta-analysis of genome-wide association studies (GWAS) involving 1,654,960 participants from various ancestries that uncovered 773 genomic loci associated with at least one lipid trait from standard lipid profiling (i.e. high-density lipoprotein cholesterol [HDL-C], LDL-C, triglycerides [TG] and total cholesterol [Total C]) in one or more ancestries (13).

From GWAS data, it is possible to aggregate polygenic effects into a single metric to predict the risk of complex diseases. Several methods to calculate these polygenic risk scores (PRS) are now available (14). These instruments have shown promising predictive performance for several diseases including CAD (15). When added to traditional risk factors and clinical scores, PRS improve the predictive performance for myocardial infarction (MI), especially in middle-aged men (16,17). Implementing these tools in clinical settings has the potential to delay the development of CAD with appropriate interventions in patients at high genetic risk (18–20).

In this study, we combined PRS for CAD and several circulating lipoproteins and fatty acids to predict the incidence of CAD, defined as MI or revascularization procedures, in UK Biobank (UKB). Our main objectives were to: (i) perform genome-wide association analyses on circulating blood lipids measured by NMR in individuals without lipid-lowering treatments, (ii) derive PRS and evaluate their association with the incidence of CAD, and (iii) combine all independent PRS to create a multivariable model and evaluate its performance to predict incident CAD.

## Methods

Data that support our findings are available through appropriate application to UK Biobank (UKB). UKB is a large-scale prospective study that received approval from the British National Health Service, North West - Haydock Research Ethics Committee (16/NW/0274). This cohort includes electronic health records of > 500,000 individuals aged between 40 and 73 years at baseline and recruited from 2006 to 2010 in the United Kingdom. All participants of UKB provided informed consent at the baseline assessment. Data access permission for this study was granted under UKB application 25205.

Full detailed methods of this study are available in the supplemental materials. Computational codes are available, without compromising sensitive individual-level data, from the corresponding author upon reasonable request. Summary statistics of genome-wide association studies and effect sizes to calculate PRS will be deposited respectively on GWAS Catalog platform (https://www.ebi.ac.uk/gwas/) and PGS Catalog (https://www.pgscatalog.org).

## Results

Supplemental Table I presents the baseline characteristics of UK Biobank participants included in each analysis. GWAS were performed for 29 blood lipid traits in 73,915 individuals not under lipid-lowering treatment (Figure 1). A total of 16,531,032 SNPs were screened during the process. Supplemental Table II reports the number of genome-wide associated loci for each trait, ranging between 19 and 59. Estimated narrow-sense heritability varied between 18% and 28%. The number of genes identified using the MAGMA algorithm implemented in FUMA ranged between 88 (docosahexaenoic acid, DHA) and 240 (ApoB and Omega 6) (Supplemental Table II). The number of common genes between the GWAS for CAD and blood lipids traits from NMR ranged between 0 (HDL-P) and 11 (ApoB, ApoB / apolipoprotein A1 [ApoA1], small LDL particles [S LDL-P], LDL particles [LDL-P], VLDL cholesterol [VLDL-C], VLDL particles [VLDL-P], remnant cholesterol [Remnant C]), representing 6.6% or less of the genes per blood lipid trait. Of the 44 genes identified from CARDIoGRAMplusC4D, 12 (27%) were also identified in at least one or more NMR blood lipids GWAS and 24 (55%) are common with at least one trait from the latest GLGC GWAS. Between 83% and 100% of the genes identified in the NMR lipoprotein GWAS were common with one of the standard lipid traits included in the GLGC GWAS, while this proportion varied from 67% to 97% for the NMR FA GWAS (Supplemental Table II). The mapped genes for the lipoprotein traits could all be found in previous GWAS of lipid traits, except for four genes mapped from the IDL-C, IDL-P or XL-VLDL-C GWAS (Supplemental Table III).

**Figure 1:**
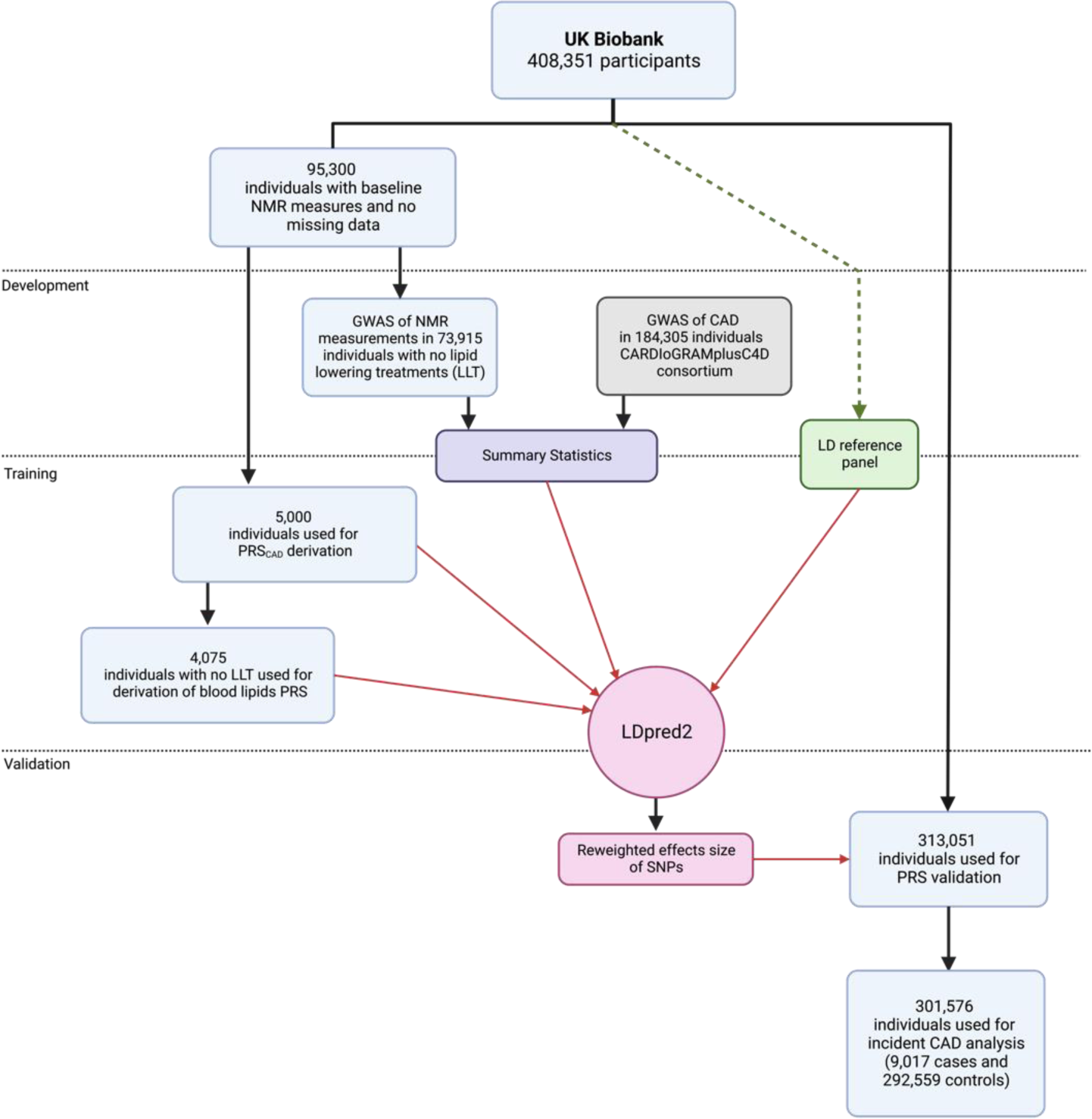
Flowchart of the study design. Overview of participants’ selection for genome-wide association analyses (i.e. development) and PRS calculation (i.e. training and validation). CAD: coronary artery disease; LD: linkage disequilibrium; LLT: lipid-lowering treatments; NMR: nuclear magnetic resonance; PRS: polygenic risk score; SNPs: single nucleotide polymorphisms

PRS were derived in the training dataset for each blood lipid trait (n=4,075 individuals, 1,114,806 SNPs) and CAD (n=5,000 individuals, 1,111,015 SNPs) (Supplemental Table IV) and calculated in the validation dataset (n=313,051 individuals). In the training dataset (n=4,075), the lipid PRS all remained strongly associated with their respective lipid trait despite adjustment for PRS_CAD_ (Supplemental Table V). As shown in the correlogram (Figure 2), PRS_CAD_ was either weakly or not correlated with all other blood lipids PRS (absolute Pearson’s correlation r coefficient between 0.01 and 0.19). Using hierarchical clustering, we identified eleven clusters in which PRS were highly correlated with each other (absolute r≥0.71, Supplemental Table VI and Figure 2). There were four clusters for lipoprotein PRS and 6 clusters for FA PRS (Supplemental Table VI and Figure 2).

**Figure 2:**
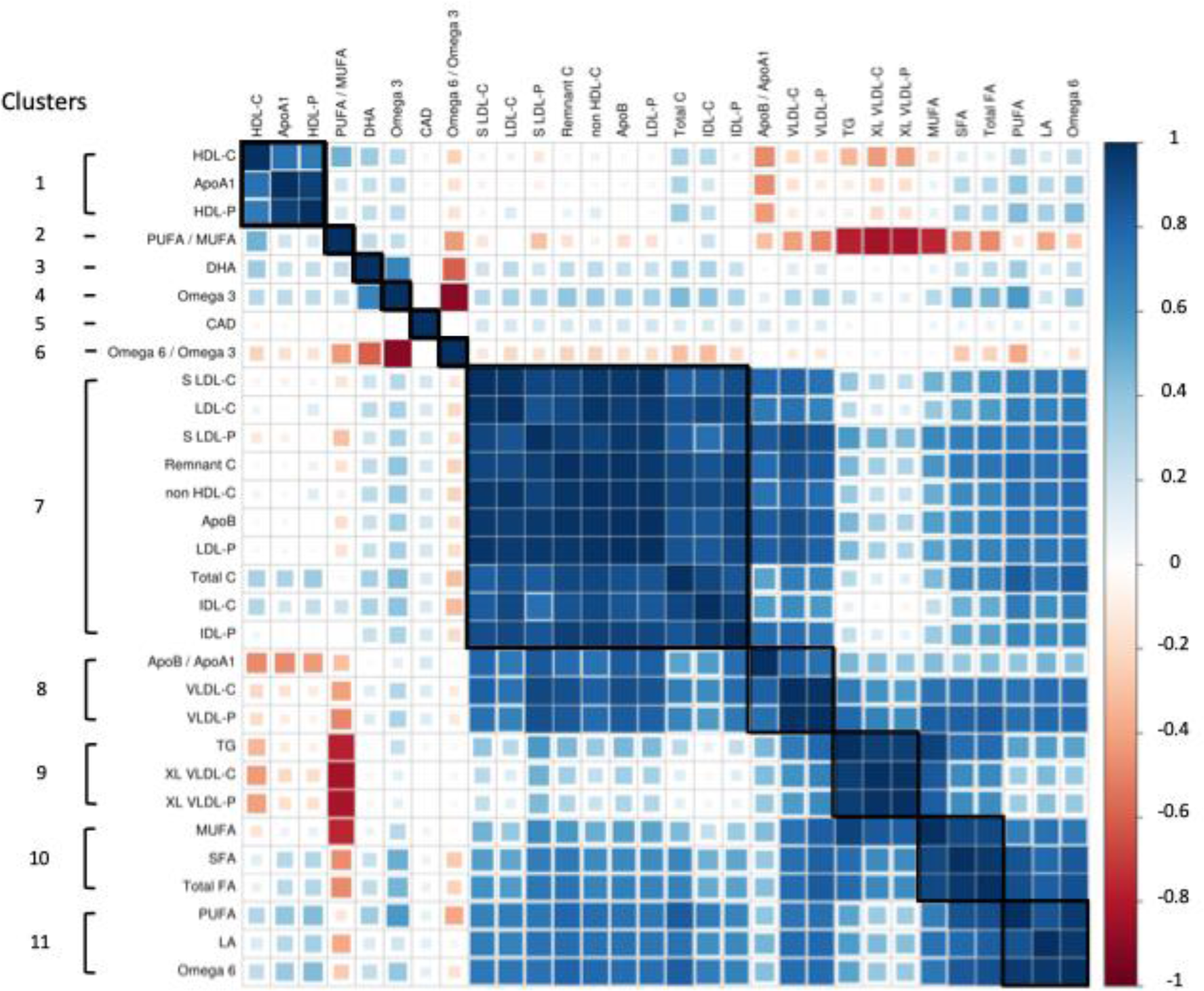
Correlation between PRS for circulating blood lipids measured by NMR and PRS_CAD_. Data are presented as gradient of Pearson’s coefficient of correlation (r) in the validation dataset (n = 313,051) and clusters of correlated PRS. ApoA: apolipoprotein A1; ApoB: apolipoprotein B; C: cholesterol; CAD: coronary artery disease; DHA: docosahexaenoic acid; FA: fatty acids; HDL: high-density lipoprotein; IDL: intermediate-density lipoprotein; LA: linoleic acid; LDL: low-density lipoprotein; MUFA: monounsaturated fatty acids; P: Particles; PUFA: polyunsaturated fatty acids; SFA: saturated fatty acids; S LDL: small low-density lipoprotein; VLDL: very-low-density lipoprotein; Total C: total cholesterol; TG: triglycerides; XL VLDL: extra-large very low-density lipoprotein or chylomicron

We investigated the association of each PRS with CAD incidence using Cox proportional hazards models. Out of 30 PRS, 28 were associated with the incidence of CAD in the validation dataset (9,017 incident CAD out of a total of 301,576 participants, median follow-up time: 12.61 years) (Figure 3). PRS_CAD_ showed the strongest association (HR=1.59 [1.55 - 1.62], *P*<1e-300). PRS_VLDL-C_ was the most strongly associated lipoprotein PRS (HR=1.19 [1.17 - 1.22], *P*=8.85e-62) and PRS_Omega 6_ was the most strongly associated FA PRS (HR=1.13 [1.10 - 1.15], *P*=1.27e-29). PRS_DHA_ and PRS_Omega6/ Omega 3_ were not significantly associated with CAD incidence. Observational associations between the measured traits and incident CAD were concordant with these associations (Supplemental Table VII).

**Figure 3:**
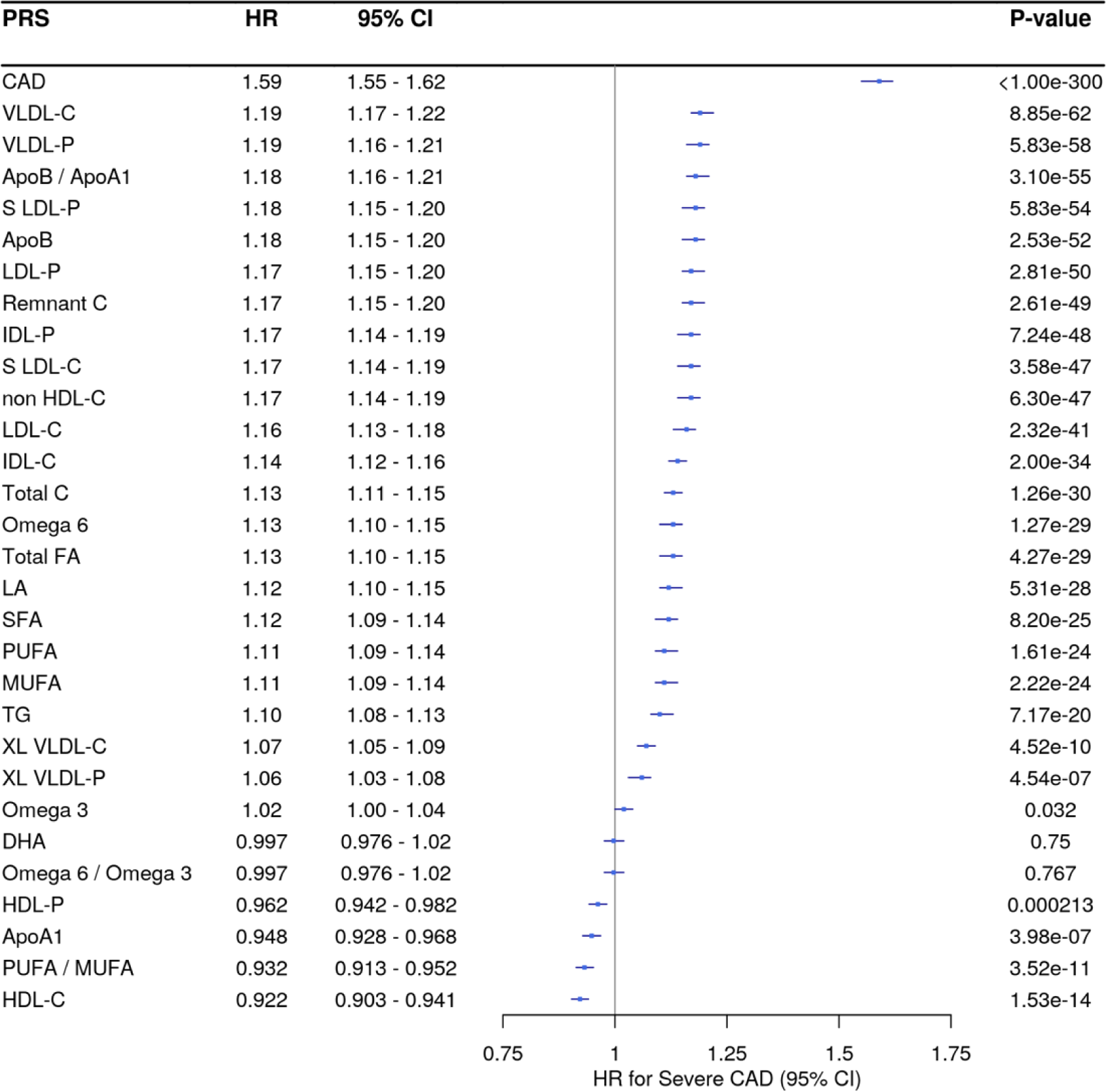
Association of PRS for blood lipids measured by NMR and PRS_CAD_ with the incidence of CAD. Cox regression results were obtained from models adjusted for age, sex and the first ten genetic principal components in all individuals without CAD at recruitment (n=301,576). Data are presented as estimated hazard ratios (HR) for incident CAD per SD increase of PRS. 95% confidence intervals (95% CI) were calculated using bootstrap (1000 iterations). ApoA1: apolipoprotein A1; ApoB: apolipoprotein B; C: cholesterol; CAD: coronary artery disease; DHA: docosahexaenoic acid; FA: fatty acids; HDL: high-density lipoprotein; IDL: intermediate-density lipoprotein; LA: linoleic acid; LDL: low-density lipoprotein; MUFA: monounsaturated fatty acids; P: Particles; PUFA: polyunsaturated fatty acids; SFA: saturated fatty acids; S LDL: small low-density lipoprotein; VLDL: very low-density lipoprotein; Total C: total cholesterol; TG: triglycerides; XL VLDL: extra-large very low-density lipoprotein or chylomicron

Following an iterative process to select independent PRS associated with CAD, four PRS were included into a multivariate model: PRS_CAD_, PRS_VLDL-C_, PRS_Total C_, and PRS_TG_ (Supplemental Table VIII). Other PRS were either correlated or did not improve the capacity of the model to identify prevalent CAD. For instance, when FA PRS were added to a model including PRS_VLDL-C_, their effect on prevalent CAD was in the opposite direction and attenuated (Supplemental Table IX).

We tested the model combining PRS_CAD_ with selected blood lipids PRS derived from NMR data in a multivariate Cox regression model, including age, sex, and PCs as covariates. Compared to the model without PRS (C-statistic=0.716 [0.711 - 0.721]), both models including PRS_CAD_ only and PRS_CAD_ with selected blood lipids PRS had significantly higher discriminative capacity with respective C-statistics reaching 0.747 [0.743 - 0.752] and 0.749 [0.744 - 0.754] (Table 1 and Figure 4A). The difference in C-statistics (Table 1) between the model with and without PRS_CAD_ was 0.031 [0.028 - 0.034]. The difference in C-statistics was similar when blood lipids PRS were added, compared to the model without PRS (0.033 [0.030 - 0.035]) (Table 1 and Figure 4B), but the difference with the model including only PRS_CAD_ was significantly different from the null (0.002 [0.001-0.003]). A likelihood ratio test (LRT) showed that the full model (i.e. including PRS_CAD_ and blood lipids PRS) significantly improved goodness of fit (chi-square test statistic=119.19, *P*=1.16e-25), compared to the nested model (i.e. including PRS_CAD_ only). When traditional risk factors were added (i.e., BMI, SBP, hypertension, diabetes, smoking, TG and LDL-C), the discriminative capacities of the model including only PRS_CAD_ (C-statistic=0.778 [0.773 - 0.783]) and the model including PRS_CAD_ with selected blood lipids PRS (C-statistic=0.778 [0.773 - 0.783]) were significantly improved when compared with the model with only risk factors (C-statistic=0.755 [0.751 - 0.760]) (Table 1 and Figure 4A). The increase in C-statistics reached 0.022 (0.020 - 0.025) for both models (Table 1 and Figure 4B). While the difference in C-statistic between the two models (with versus without blood lipids PRS) was not significantly different from the null (0 [0 - 0.001]), the model including blood lipids PRS slightly outperformed the model with only PRS_CAD_ in terms of goodness of fit (chi-square test statistic=20.18, *P*=1.56e-04).

**Figure 4:**
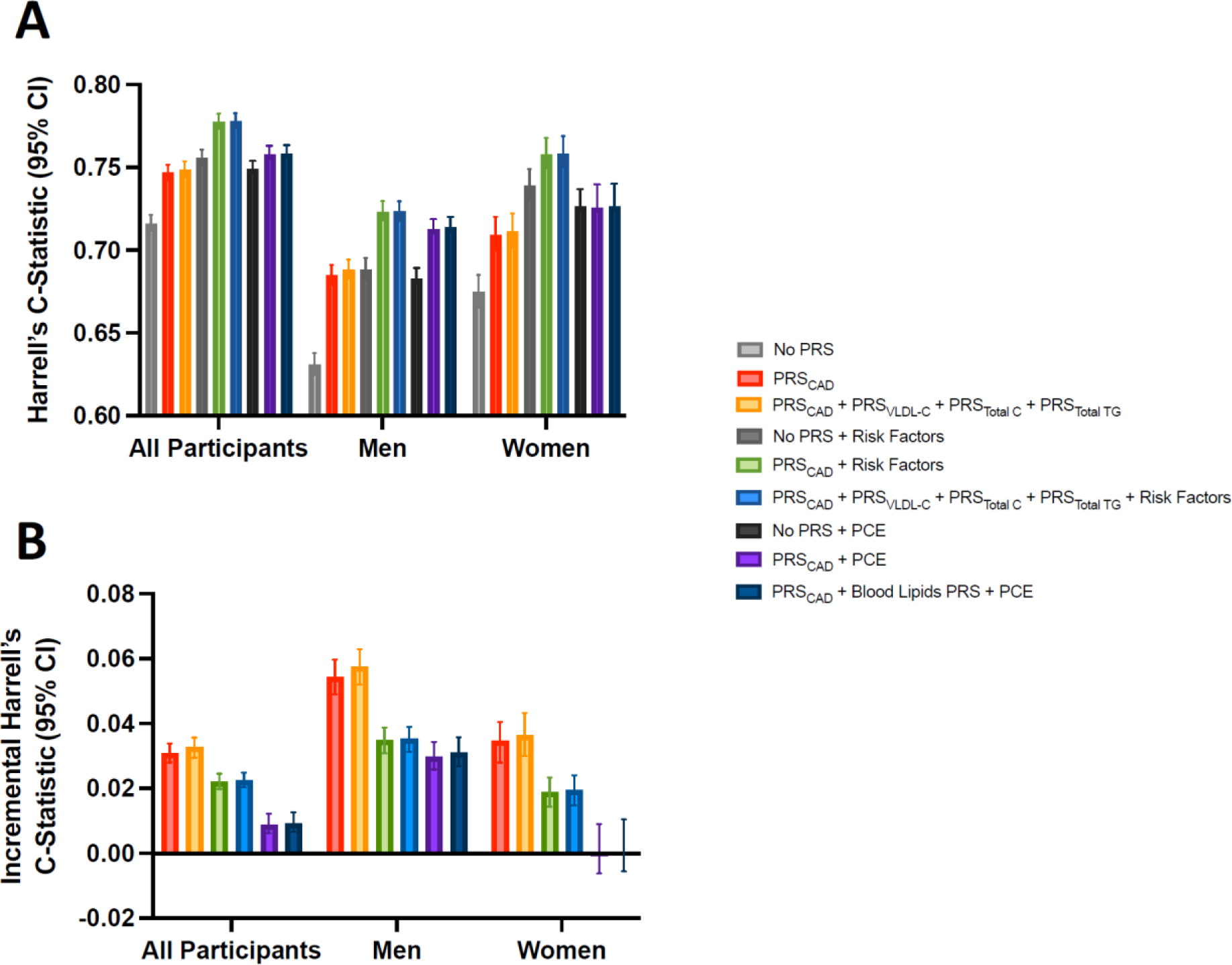
Increment in discrimination capacities using Harrell’s C-statistic in Cox regression models for the incidence of CAD. Data are presented as C-statistic (A) and increment in C-Statistic compared to the model without PRS (i.e. difference of C-statistics) (B), in all individuals, and separately in men and women. 95% confidence intervals (95% CI) were calculated using bootstrap (1000 iterations). All models include age, sex (not for sex-stratified analysis) and the first ten genetic principal components. Risk factors include body mass index, systolic blood pressure, hypertension, diabetes, smoking status, triglycerides levels and LDL-C levels. C: cholesterol; CAD: coronary artery disease; P: Particles; PCE: Pooled Cohort Equation; PRS: polygenic risk score; VLDL: very low-density lipoprotein; Total C: total cholesterol; TG: triglycerides.

**Table 1:** Comparison of discriminative capacities using Harrell’s C statistic in Cox regression models for the incidence of CAD. Cox regression models adjusted for age, sex and the first ten genetic principal components (PCs) in all individuals without a previous history of CAD (n=301,576), and only age and PCs for sex-stratified analyses. Difference between the C-statistic (Delta C-statistic) of the studied model and the C-statistic of the model without PRS or with PRS_CAD_ only are reported. Risk factors include body mass index, systolic blood pressure, hypertension, diabetes, smoking status, triglycerides levels and low-density lipoprotein cholesterol levels (corrected if lipid lowering treatments were taken). 95% confidence intervals were obtained by bootstrapping. C: Cholesterol; CAD: coronary artery disease; NA: not applicable; VLDL: very low-density lipoprotein; TG: triglycerides.

| Sex | Model | Cases (n) | Total (n) | C-statistic (Bootstrap 95%CI) | Increment in model's C-statistic compared to the model without PRS (Bootstrap 95%CI) | Increment in model's C-statistic compared to the model equivalent model including PRS<sub>CAD</sub> only (Bootstrap 95%CI) |
| --- | --- | --- | --- | --- | --- | --- |
| All | No PRS | 9,017 | 301,576 | 0.716 (0.711 to 0.721) | NA (NA) | NA (NA) |
|  | PRS<sub>CAD</sub> | 9,017 | 301,576 | 0.747 (0.742 to 0.751) | 0.031 (0.028 to 0.034) | NA (NA) |
|  | PRS<sub>CAD</sub> + PRS<sub>VLDL-C</sub> + PRS<sub>Total C</sub> + PRS<sub>TG</sub> | 9,017 | 301,576 | 0.749 (0.744 to 0.753) | 0.033 (0.030 to 0.036) | 0.002 (0.001 to 0.003) |
|  | No PRS + Risk Factors | 7,990 | 269,090 | 0.755 (0.751 to 0.760) | NA (NA) | NA (NA) |
|  | PRS<sub>CAD</sub> + Risk Factors | 7,990 | 269,090 | 0.778 (0.773 to 0.782) | 0.022 (0.020 to 0.025) | NA (NA) |
|  | PRS<sub>CAD</sub> + PRS<sub>VLDL-C</sub> + PRS<sub>Total C</sub> + PRS<sub>TG</sub> + Risk Factors | 7,990 | 269,090 | 0.778 (0.773 to 0.783) | 0.022 (0.020 to 0.025) | 0 (0 to 0.001) |
|  | No PRS + PCE | 7,814 | 266,530 | 0.749 (0.743 to 0.754) | NA (NA) | NA (NA) |
|  | PRS<sub>CAD</sub> + PCE | 7,814 | 266,530 | 0.758 (0.753 to 0.763) | 0.009 (0.006 to 0.012) | NA (NA) |
|  | PRS<sub>CAD</sub> + PRS<sub>VLDL-C</sub> + PRS<sub>Total C</sub> + PRS<sub>TG</sub> + PCE | 7,814 | 266,530 | 0.758 (0.754 to 0.763) | 0.009 (0.007 to 0.012) | 0 (0 to 0.001) |
| Men | No PRS | 6,491 | 134,713 | 0.631 (0.625 to 0.638) | NA (NA) | NA (NA) |
|  | PRS<sub>CAD</sub> | 6,491 | 134,713 | 0.685 (0.679 to 0.691) | 0.054 (0.049 to 0.059) | NA (NA) |
|  | PRS<sub>CAD</sub> + PRS<sub>VLDL-C</sub> + PRS<sub>Total C</sub> + PRS<sub>TG</sub> | 6,491 | 134,713 | 0.688 (0.683 to 0.695) | 0.058 (0.052 to 0.063) | 0.003 (0.002 to 0.005) |
|  | No PRS + Risk Factors | 5,775 | 120,414 | 0.688 (0.682 to 0.695) | NA (NA) | NA (NA) |
|  | PRS<sub>CAD</sub> + Risk Factors | 5,775 | 120,414 | 0.723 (0.718 to 0.730) | 0.035 (0.031 to 0.039) | NA (NA) |
|  | PRS<sub>CAD</sub> + PRS<sub>VLDL-C</sub> + PRS<sub>Total C</sub> + PRS<sub>TG</sub> + Risk Factors | 5,775 | 120,414 | 0.723 (0.718 to 0.730) | 0.035 (0.031 to 0.040) | 0 (0 to 0.001) |
|  | No PRS + PCE | 5,649 | 118,876 | 0.683 (0.676 to 0.689) | NA (NA) | NA (NA) |
|  | PRS<sub>CAD</sub> + PCE | 5,649 | 118,876 | 0.712 (0.707 to 0.718) | 0.03 (0.026 to 0.034) | NA (NA) |
|  | PRS<sub>CAD</sub> + PRS<sub>VLDL-C</sub> + PRS<sub>Total C</sub> + PRS<sub>TG</sub> + PCE | 5,649 | 118,876 | 0.714 (0.707 to 0.720) | 0.031 (0.027 to 0.036) | 0.001 (0.001 to 0.002) |
| Women | No PRS | 2,526 | 166,863 | 0.675 (0.666 to 0.686) | NA (NA) | NA (NA) |
|  | PRS<sub>CAD</sub> | 2,526 | 166,863 | 0.709 (0.701 to 0.720) | 0.035 (0.028 to 0.041) | NA (NA) |
|  | PRS<sub>CAD</sub> + PRS<sub>VLDL-C</sub> + PRS<sub>Total C</sub> + PRS<sub>TG</sub> | 2,526 | 166,863 | 0.711 (0.702 to 0.722) | 0.036 (0.030 to 0.043) | 0.002 (0.001 to 0.004) |
|  | No PRS + Risk Factors | 2,215 | 148,676 | 0.739 (0.730 to 0.748) | NA (NA) | NA (NA) |
|  | PRS<sub>CAD</sub> + Risk Factors | 2,215 | 148,676 | 0.758 (0.749 to 0.768) | 0.019 (0.015 to 0.023) | NA (NA) |
|  | PRS<sub>CAD</sub> + PRS<sub>VLDL-C</sub> + PRS<sub>Total C</sub> + PRS<sub>TG</sub> + Risk Factors | 2,215 | 148,676 | 0.758 (0.750 to 0.769) | 0.020 (0.015 to 0.024) | 0.001 (0 to 0.002) |
|  | No PRS + PCE | 2,165 | 147,654 | 0.726 (0.715 to 0.736) | NA (NA) | NA (NA) |
|  | PRS<sub>CAD</sub> + PCE | 2,165 | 147,654 | 0.726 (0.717 to 0.739) | -0.001 (-0.007 to 0.010) | NA (NA) |
|  | PRS<sub>CAD</sub> + PRS<sub>VLDL-C</sub> + PRS<sub>Total C</sub> + PRS<sub>TG</sub> + PCE | 2,165 | 147,654 | 0.726 (0.717 to 0.740) | 0 (-0.006 to 0.011) | 0.001 (0 to 0.002) |

Similarly, when the PCE was used, the discriminative capacities of the model including only PRS_CAD_ (0.758 [0.753 - 0.763]) and the model including PRS_CAD_ with selected blood lipids PRS (C-statistic=0.758 [0.754 - 0.763]) were significantly improved when compared to the model without PRS (C-statistic=0.749 [0.743 - 0.754]) (Table 1). The difference in C-statistic between the two models (with versus without blood lipids PRS) was not different from the null (0 [0 - 0.001]), although the model including blood lipids PRS slightly outperformed the model with only PRS_CAD_ in terms of goodness of fit (chi-square test statistic=39.58, *P*=1.31e-08).

Similar results were obtained when analyses were stratified by sex (Table 1 and Figure 4).

Categorized net reclassification improvement (NRI) was calculated using a threshold of 3% (NRI^0.03^) of predicted 10-year risk of CAD. The addition of PRS to the Cox proportional hazard models including PCE resulted in a NRI^0.03^ of 0.059 (95% CI, [0.047 to 0.069]), mostly driven by the up reclassification of cases. Continuous NRI (NRI^>0^) was equal to 0.374 (95% CI [0.344 to 0.401]) (Supplemental Table X). In age-stratified analysis, individuals from the first tertile of the age distribution (i.e., individuals aged between 40 and 54 years) show the highest reclassification performance with an overall NRI^0.03^ reaching 0.135 (95% CI [0.110 to 0.173]), while NRI^>0^ reached 0.508 (95% CI [0.446 to 0.564]) (Supplemental Table X).

## Discussion

We developed PRS from blood lipids measured using NMR in 73,915 European ancestry participants from UKB. Among all tested PRS, 27 were associated with the incidence of CAD in a middle-aged white British population. However, combining a PRS_CAD_ with several independent blood lipids PRS offered marginal improvement in discriminative capacity over a PRS_CAD_ alone.

Our genome-wide association analyses on 29 circulating blood lipids traits measured using high-throughput NMR spectroscopy confirmed that these traits involve multiple genomic loci and have strong heritabilities (21). A high proportion of the genes identified from these GWAS were already reported in a recently published GWAS for standard lipid traits from the GLGC (13). However, only a few were retrieved in the CAD GWAS from CARDIoGRAMplusC4D (22), consistent with the weak correlation observed between PRS_CAD_ and blood lipids PRS. Using an independent subset of UKB, we found that out of 29 circulating blood lipids PRS, 27 were associated with the incidence of CAD, including all tested lipoproteins PRS. This adds more lipoprotein-related traits to the previously determined list of PRS associated with CAD in British adults (23) or with the extent of CAD in Icelandic adults (i.e. HDL-C, LDL-C, non-HDL-C and TG) (24). More specifically, in our study, PRS for particles and cholesterol content of ApoB-containing lipoproteins (i.e. small LDL, LDL, IDL, VLDL, remnant and chylomicron) predicted similarly the incidence of CAD. Globally, these associations were significantly stronger than total C or total TG. VLDL-C and VLDL-P were the most associated lipoproteins PRS with the incidence of CAD, although they were strongly correlated with genetically-predicted levels of ApoB (Pearson’s correlation r=0.88 for VLDL-C and 0.84 for VLDL-P). The results are consistent with the strong association between VLDL-C measured with NMR in participants without lipid-lowering treatment in UKB and incident myocardial infarction observed previously (8).(6)(4) These findings further emphasize the importance of VLDL particles, which have been shown to be trapped preferentially within the intima in animal models (25).

We also provide new insights concerning the association between circulating FA and the incidence of CAD. In our analysis, the most associated FA PRS were PRS_Omega-6_ and PRS_Total FA_ (positive association). Significant associations were also observed for PRS for PUFA, mono-unsaturated fatty acids (MUFA), linoleic acid (LA) and saturated fatty acids (SFA). However, the PRS for all these FA traits were highly correlated to ApoB containing lipoproteins PRS, notably to PRS_VLDL-C_ and PRS_VLDL-P_ (with Pearson’s correlation r>0.7). Adding PRS_VLDL-C_ in the models inverted and attenuated these associations in multivariate analyses. We also show that the PRS_PUFA / MUFA_ is inversely associated with genetically-predicted TG and TG-rich lipoproteins such as chylomicrons as well as the incidence of CAD. Previous observational studies showed an overall protective role of circulating PUFA measured by NMR, whereas MUFA levels were associated with an increased cadiovascular risk (8,26). However, a recent Mendelian randomization study using the same NMR data in UKB did not find convincing evidence of a protective role for circulating PUFA on the risk of cardiovascular diseases. The authors of this study suggested a possible bias caused by horizontal pleiotropy, notably due to the presence of variants at the *FADS* locus implicated in lipoprotein metabolism in their genetic instruments (27). In this sense, we also report here a high number of common genes between FA GWAS and lipid profiles GWAS. Pleiotropic effects of variants involved in both FA and lipoprotein metabolism make it difficult to draw conclusions concerning the protective or deleterious role of circulating PUFA. Finally, PRS of specific types of PUFA, such as PRS_DHA_, PRS_Omega 6 / Omega 3_ and PRS_Omega 3_ were not or weakly associated with the incidence of CAD. Circulating total omega-3 FA and specific omega-3 FA fractions, including DHA and eicosapentaenoic acid (EPA) levels were already reported as conferring a lower cardiovascular risk and associated to a lower mortality rate when naturally found at high levels in blood (28). However, recent evidence suggests that EPA might drive the cardioprotective role, as the combination of EPA and DHA has not shown convincing benefits in clinical trials (29). In contrast, high doses of icosapent ethyl, a purified EPA ethyl ester, were shown to reduce TG plasma levels and cardiovascular events in high-risk individuals with hypertriglyceridemia already on statin therapy(11,29,30). Of note, we could not develop a PRS for EPA since EPA levels were not available in UKB.

Overall, despite the fact that most circulating blood lipids PRS individually predicted the incidence of CAD, adding multiple independent blood lipids PRS in a model already including PRS_CAD_ increased only marginally the prediction of CAD. One explanation for this result could be that the PRS_CAD_ already captures part of the genetic risk linked to blood lipids. In our study, out of the 44 significant genes identified from the meta-analysis of CARDIoGRAMplusC4D (22), 12 genes (27%) were found in common with our GWAS of circulating blood lipids, whereas 24 genes (55%) were common to at least one of the four lipid traits included in the latest GLGC GWAS (13). The lipid PRS however remained strongly associated with the corresponding lipid trait after adjusting for PRS_CAD_. A previous study led in a Finnish population evaluated the association of a combination of PRS for LDL-C, TG and CAD with myocardial infarction or revascularization. They showed that the associations for the two lipid PRS were slightly attenuated when adusting for PRS_CAD_, but remained significant, with OR of 1.11 and 1.06 per SD, respectively, in the complete model (31).

Furthermore, we used data from a population of middle-aged individuals with a low event rate. A substantial proportion (15.1%) were already on lipid-lowering treatments at baseline, without having a prior history of CAD. A possible selection bias related to the healthier lifestyle of UKB participants compared to the UK general population was already reported (32). Lipid-lowering treatment and a healthy lifestyle could protect against high genetically-determined lipid levels.

Combining PRS for more risk factors could potentially further improve risk prediction. For instance, Ramirez et al. have combined PRS for common risk factors associated with CAD (i.e. BMI, SBP, diastolic blood pressure, C-reactive protein, type 2 diabetes, heart failure, standard HDL-C, standard LDL-C, standard TG, T-peak-to-T end interval) using data from UKB. They improved significantly their models’ discriminative capacities when PRS for risk factors were added to a PRS_CAD_. However, the PRS for risk factors did not significantly improve the models’ discriminative capacities when considering also the QRISK3 clinical risk score (23). Nonetheless, genetic information to calculate PRS for all traits is available with genome-wide genotyping once in a lifetime. We show statistically significant improvement in reclassification when adding multiple PRS, including a PRS_CAD_, to a clinical risk score, as shown previously (17). Blood lipids and other risk factors PRS could also help to identify at a young age individuals presenting higher genetic susceptibilities, providing an opportunity for early preventive interventions. Notably, PRS have been shown to perform better to predict early-onset events (16).

Our study has a few limitations. First, the blood lipids PRS that we derived combined the effect of common SNPs, and thus did not include rare monogenic variants that are responsible for severe dyslipidemias. Second, our analysis was restricted to individuals from white British ancestry. Several other ethnic backgrounds, such as South Asians, are more susceptible to CAD and MI (33,34). The utility of blood lipids PRS needs to be evaluated in other ancestries. Third, we mostly assessed the predictive performance using the Harrell’s C-statistic. It has already been demonstrated that this metric is not a perfect indicator of the added value conferred by new predictors in a same model (35). This is why we also compared goodness of fit using the likelihood ratio test. Fourth, we used a modest number of individuals in our GWAS compared to large consortia meta-analyses. However, our study design allowed us to derive PRS for well-characterized blood lipid traits derived from NMR and to avoid sample overlap. The addition of other independent cohorts, including younger individuals and other ancestries, has the potential to increase the discriminative capacities of PRS. Fifth, the derived PRS should be validated and tested in other external and independent cohorts.

In conclusion, PRS for circulating lipoproteins and fatty acids are associated with the incidence of CAD. However, combining selected independent blood lipids PRS provided a marginal improvement in the discriminative capacity provided by the PRS_CAD_ alone. Further studies are needed to evaluate the impact of preventive interventions in individuals at high genetic risk.

## Data Availability

Computational codes are available, without compromising sensitive individual-level data, from the corresponding author upon reasonable request. Summary statistics of genome-wide association studies and effect sizes to calculate PRS will be deposited respectively on GWAS Catalog platform (https://www.ebi.ac.uk/gwas/) and PGS Catalog (https://www.pgscatalog.org).

## Non-Standard Abbreviations and Acronyms

ApoA1: Apolipoprotein A1
ApoB: Apolipoprotein B
CAD: Coronary Artery Disease
CRF: Clinical Risk Factors
DHA: Docosahexaenoic Acid
EPA: Eicosapentaenoic Acid
FA: Fatty Acids
FUMA: Functional Mapping and Annotation
GLGC: Global Lipid Genetic Consortium
GWAS: Genome Wide Association Study
HES: Hospital Episode Statistics
IDL: Intermediate Density Lipoproteins
IS: Ischemic Stroke
LA: Linoleic Acid
LD: Linkage Disequilibrium
LLT: Lipid Lowering Treatments
Lp(a): Lipoprotein(a)
MAGMA: Multi-marker Analysis of GenoMic Annotation
MI: Myocardial Infarction
MUFA: Mono-Unsaturated Fatty Acids
NMR: Nuclear Magnetic Resonance
NRI: Net Reclassification Improvement
OPSC-4: Office Of Population, Censuses and Surveys Classification of Interventions and Procedures, Version 4
PCs: First Ten Genetic Principal Components
PCE: Pooled Cohort Equation
PRS: Polygenic Risk Score
PUFA: Polyunsaturated Fatty Acids
SFA: Saturated Fatty Acids
SBP: Systolic Blood Pressure
TC: Total Cholesterol
TG: Triglycerides
UKB: UK Biobank
VLDL: Very Low-Density Lipoproteins
XL-VLDL: Extra Large Very Low-Density Lipoproteins or Chylomicrons

## Acknowledgement

We would like to thank all study participants and scientists involved in UK Biobank and in the CARDIoGRAMplusC4D Consortium.

## Supplemental Materials

Supplemental Methods

Supplemental Tables

References (8,13,16,22,36–44)

